# Toward respiratory support of critically ill COVID-19 patients using repurposed kidney hollow fiber membrane dialysers to oxygenate the blood

**DOI:** 10.1101/2020.04.06.20055236

**Authors:** David M. Rubin, Neil Stacey, Tonderayi Matambo, Claudia Do Vale, Martin J. Sussman, Tracy Snyman, Mervyn Mer, Diane Hildebrandt

**Affiliations:** Biomedical Engineering Research Group, School of Electrical Information Engineering, University of the Witwatersrand, Johannesburg, South Africa; School of Chemical and Metallurgical Engineering, University of the Witwatersrand, Johannesburg, South Africa; Institute for the Development of Energy for African Sustainability, University of South Africa; Morningside Hospital, Johannesburg; Department of Medicine, Division of Nephrology, University of the Witwatersrand, Johannesburg, South Africa; Cardio-thoracic Surgery, Milpark Hospital, Johannesburg; Department of Chemical Pathology, University of the Witwatersrand, Johannesburg; National Health Laboratory Service, Johannesburg; Charlotte Maxeke Johannesburg Academic Hospital; Department of Medicine, Divisions of Critical Care and Pulmonology, University of the Witwatersrand, Johannesburg

**Keywords:** Blood oxygenator, repurposing of renal dialysis, oxygen transfer characteristics

## Abstract

The COVID-19 pandemic has highlighted resource constraints in respiratory support. The oxygen transfer characteristics of a hollow fiber membrane dialyser were investigated with a view to repurposing the device as a low-cost, readily available blood oxygenator. Oxygen transfer in a low-flux hollow fiber dialyser with a polysulfone membrane was studied by passing first water and then blood through the dialyser in counter-current to high-purity oxygen. Oxygen transfer rates of about 15% of the nominal 250 *ml*(*STP*)/*min* of a typical adult oxygen consumption rate were achieved for blood flow rates of 500*ml/min*. Using two such dialysis devices in parallel could provide up to 30% of the nominal oxygen consumption. Specific hollow fiber dialysis devices operating with suitable pumps in a veno-venous access configuration, could provide a cost-effective and readily available supplementation of respiratory support in the face of severe resource constraints.

## I. INTRODUCTION

The COVID-19 pandemic has highlighted resource constraints in the management of respiratory distress [1], and debate continues on the merits of ventilation v.s. less invasive respiratory support for COVID-19, in terms of disease management and safety of medical staff [2]. Extracorporeal membrane oxygenators (ECMO) have been used successfully to oxygenate and decarbonate blood [3], [4], and there are a number of indications for the use of ECMO [5]. However, its use is costly and resource intensive [6].

Our objective is not to replace ECMO but rather to investigate repurposing of renal hollow fiber membrane dialysers (HFMDs) as cost-effective augmentation of respiratory support in resource constrained environments.

Unlike purpose-designed ECMO membranes [7], [8], renal dialysis membranes are designed for liquid-liquid transfer of specific molecules. Nonetheless, their existing clinical approval for renal replacement therapy makes repurposing HFMDs as oxygenators an attractive option.

Nominal *O*_2_ consumption in adults is approximately 250 *ml · min*^−1^*(STP)* (0.357*g · min*^−1^) [9]. The purpose of respiratory support is to achieve concentrations of *O*_2_ and *CO*_2_ in arterial blood that are compatible with proper organ function. Repurposed dialysers could potentially provide sufficient gas transfer to compensate for the respiratory deficit in some patients.

We investigated *O*_2_ transfer in a renal HFMD using water as a blood substitute. This facilitated exclusion of problems such as air bubble formation and trans-membrane fluid leakage. The minimum *O*_2_ transfer rate in water motivated further experiments with blood.

## II. MATERIALS AND METHODS

### A. Theoretical considerations and computational methodology

Mass transfer in the HFMD is modelled by the following equation:

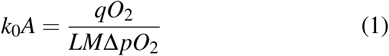

where, *k*_0_ is the mass transfer coefficient, *A* is the membrane area, *qO*_2_ is the rate of oxygen transferred and *LM*Δ*pO*_2_ is the logarithmic mean of partial pressure differences at the top and bottom of the device.

Defining the partial pressure difference between the blood inlet and the oxygen flow outlet as 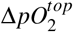, and between the blood outlet and oxygen flow inlet as 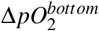, and expanding the definition of logarithm mean, the expression for the mass transfer coefficient × Area *(k*_0_*A)* becomes:

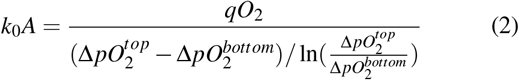

To achieve a given *qO*_2_, this equation facilitates determination of the minimum *k*_0_*A* of the device.

As oxygen transport capacity of blood resides primarily in haemoglobin, it reaches saturation at a low partial pressure of oxygen. Thus we can assume that oxygen transfer into blood takes place within a narrow range of partial pressures. Also, for high-purity oxygen, the *pO*_2_ may be regarded as essentially constant.

The oxygen carrying capacity of blood is 8800*μM* of *O*_2_ bound to the Hb for 100% saturation [10] and dissolved *O*_2_ in blood is negligible. The HFMD cartridges are designed for blood flow-rates up to 500 *ml/min*, and we evaluate oxygen-carrying capacity of blood for a Hb concentration of 15*g*/100*ml* and a typical oxyhaemoglobin dissociation curve.

At sea level atmospheric pressure, and for a typical venous and arterial *pO*_2_ of 6 and 12.7 *kPa* (and corresponding *HbSat* of 45% and 95%) respectively, the oxygen uptake in 500 *ml* of blood and the 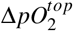 and 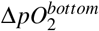 are calculated and substituted into (2) to estimate the required *k*_0_*A*. This is repeated for an inlet venous *HbSat* of 25% as shown in Table I.

**TABLE I.** Theoretical calculated maximum oxygen uptake per 500 ml of blood and required *k*_0_*A* for specified inlet conditions where the outlet is near saturation with *pO*_2_ of 95mmHg.

| Inlet<br>$PO_2mmHg(HbSat)$ | Max $O_2$ uptake<br>$mg/500ml$ blood | $k_0A$<br>$mg \cdot min^{-1} \cdot kPa^{-1}$ |
| --- | --- | --- |
| 45 (76%) | 35.7 | 0.389 |
| 25 (45%) | 73.4 | 0.787 |

It is assumed that mass transfer across the membrane is rate limiting rather than blood mass transfer or the reaction of oxygen with haemoglobin.

### B. Materials and methods for gas transfer into water

The experiments were conducted at the Biotechnology Laboratory at UNISA, Johannesburg at an altitude of about 1700*m* with atmospheric pressure about 84*kPa* and temperature about 25*^o^C*.

A low-flux Leoceed-21N (Asahi Kasei Medical) hollow fiber membrane dialyser unit renal dialysis cartridge was used in this study. The polysulfone membrane has an effective surface area of 2.1 m^2^, and the HFMD has a priming volume of 108 *ml*, and an internal fibre diameter and wall thickness of 185 *μm* and 35 *μm* respectively. The manufacturer-specified maximum blood flow rate is 500 *ml · min*^−1^ (mean residence time about 13*s* at this flow rate). Maximum transmembrane pressure (TMP) is rated at 80*kPa*.

Tap water was deoxygenated by boiling and allowed to cool in a sealed glass bottle, and measured for dissolved oxygen (SD 400 OXI L, Lovibond, Amesbury, UK). The deoxygenated water was pumped through the vertically mounted HFMD using a peristaltic pump (Qdos Chemical metering pumps, Watson Marlow, Cornwell, UK) via the blood inlet at 50, 200 and 500 *ml · min*^−1^ and discharged into a beaker covered with parafilm to minimize oxygen losses.

High-purity (>99%) oxygen (Afrox gas, Johannesburg, South Africa) was passed counter-current through the HFMD via the dialysate inlet, using a pressure regulator set between 60 to 80 *kPa* (gauge) followed by a needle valve to control flow rate at 400 *ml · min*^−1^.

### C. Materials and methods for gas transfer into blood

This study with human blood was approved by the institutional review board of the South African National Blood Services Human Research Ethics Committee (certificate number: 2019/0521), and the institutional review board of the University of the Witwatersrand, Johannesburg Human Research Ethics Committee (Medical) (certificate number: M200456). The experiments were conducted at the Chemical Pathology Laboratory, University of the Witwatersrand, Johannesburg, at an altitude of about 1500 *m* (atmospheric pressure about 85*kPa)*.

Recently expired donor whole blood (SANBS) was warmed in a water bath at 37*^o^C* and pooled in a glass flask to constitute approximately 4.5 *l* to which approximately 3000 units of heparin (Heparin sodium mucosal Fresenius Kabi, 5ml of 1000 *U/ml)* was added.

**Fig. 1.**
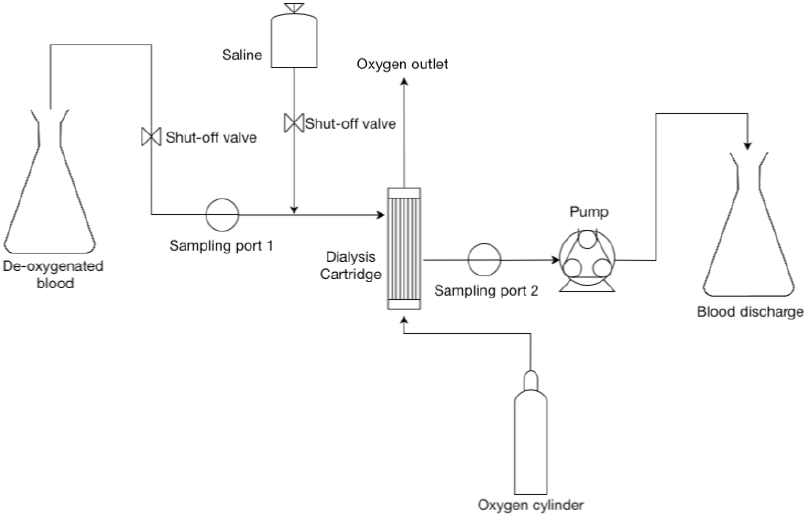
Flowsheet of experimental setup with blood. The oxygen was passed through the dialyser via the dialysate inlet while deoxygenated blood was passed through the hollow fibres in counter-current to the oxygen flow.

Nitrogen sparge gas was bubbled through the blood to reduce the % oxygen saturation of haemoglobin *(HbSat*) to typical venous levels [11]. The nitrogen flow was gradually increased from 1 *l/min* until froth was visualised,

As shown in Figure 1, blood was pumped by a roller pump through the hollow fibers of a vertically-mounted HFMD, from top to bottom, at 500 *ml/min*. The upstream placement of the roller pump reduces the risk of red blood cell lysis prior to oxygenation. Blood was discharged into a collecting flask. Oxygen was passed counter-current at 1250 *ml/min* through the outside of the fibers.

The pump was placed ≈ 0.5 *m* below the base of the HFMD to achieve a pressure head, thus reducing the required suction, making sampling easier. The blood was piped through dialysis tubing fitted with inlet and outlet sampling ports (Fresenius Kabi).

After priming with normal saline, flow was switched to blood which was mixed by swirling, prior to and during the experiments.

Two runs were performed using a Leoceed-21N (Asahi Kasei Medical) HFMD and the third run using a Leoceed-18N (effective surface areas of 1.8 *m*^2^, priming volume 96 *ml*). A high-flux Leoceed H-type dialyser was also tested but fluid leakage across the membrane into the gas side was noted, suggesting that this high-flux dialyser may not be suitable for oxygenation.

Two to four inlet and outlet blood samples of 1 - 4 *ml* each were taken for each run with heparin-coated syringes, and analysed with a blood gas analyser (Radiometer, ABL80 FLEX CO-OX) for Haemoglobin concentration (Hb), partial pressure of carbon dioxide (*pCO*_2_), partial pressure of oxygen (*pO*_2_), and haemoglobin saturation (*HbSat*). Bicarbonate ion concentrations (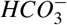) were calculated by the blood gas analyser.

Measurements were averaged for each run over the sample number. Measurement uncertainties were reported as ± 1 standard deviation based on rounded estimates from coefficients of variation [12], [13] and scaled for sample number.

## III. RESULTS

### A. Experiments with water

Oxygen was passed through the HFMD at 400 *ml/min* and water was run counter-current through the inside of the hollow fibers. Inlet oxygen concentration was measured at 3.61 *mg/l*, at 37.1C.

Outflow water was collected in beakers and oxygen concentration measured. Table II shows these data and corresponding calculated oxygen transfer rates.

**TABLE II.** Minimum oxygen transfer rates calculated from differential dissolved oxygen concentrations between inlet and outflow water.

| Water flow rate<br>($ml/min$) | Dissolved oxygen in<br>collected water<br>outflow ($mg/l$) | Calculated oxygen<br>transfer rate<br>($mg/min$) |
| --- | --- | --- |
| 50 | 28.6 | 1.25 |
| 200 | >32.9 (out of range) | >5.85 |
| 500 | >32.9 (out of range) | >14.6 |

The low reading of 28.6*mg · l*^−1^ is probably due to higher overall oxygen losses at saturation due to the slower flow rate.

Equation (2) is used to estimate *k*_0_*A* based on the minimum oxygen transfer rate of 14.6*mg · min*^−1^ at 500*ml · min*^−1^. Dissolved oxygen concentration is treated as negligible as haemoglobin carries most of the oxygen.

The oxygen exit stream is at atmospheric pressure, and the water inlet’s partial pressure is given by the % saturation read from the dissolved oxygen meter × *pO*_2_ in air at 84*kPa*, which amounts to a difference of 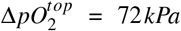. The oxygen outlet pressure has little influence on the large driving force for mass transfer.

The 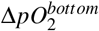 requires knowledge of the unmeasured gas pressure as it enters the HFMD. The pressure regulator reading was 60*kPa* but most of the pressure drop occurs at the needle valve. Also, prior to membrane contact there was a contractor connection and the expansion/elbow entering the membrane unit, whereas on the discharge side there was just the expansion/elbow and a short length of tubing to the discharge. While it is not possible to exactly know the pressure in the membrane unit, it must be considerably lower than the regulator’s pressure reading, and a simple estimate suggest that the relevant pressure drop is as low as 0.1*kPa*.

Because the outlet water stream is at or above saturation, the partial pressure difference at that point is at most equal to that over-pressure amount. Thus the mass transfer coefficient calculations are highly sensitive to the actual pressure in this region.

For a 0.1 *kPa* partial pressure drop across the 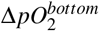, and a minimum oxygen transfer rate measured at 14.6*mg/min*, equation (2) yields a *k*_0_*A* value of 1.334 *mg · min*^−1^ *· kPa*^−1^. This implies a 3.4 fold larger oxygen mass transfer rate than required to fully oxygenate blood to 95 % saturation for incoming *pO*_2_ of 45 *mmHg*, and 1.7 fold larger than needed if the incoming *pO*_2_ is 25 *mmHg*.

Even in the unlikely case of a 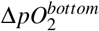 of 6*kPa*, the potential mass transfer rate of oxygen will be 1.4 fold larger than the minimum needed for incoming blood with oxygen partial pressure of 45 *mmHg* and 70% of the required amount for an incoming saturation of 25 *mmHg*.

### B. Results for gas transfer in blood

As seen in figure 2, the dark blood entering the top and bright red blood exiting from the bottom of the HFMD is clear evidence of oxygenation.

Measurements from the inlet and outlet ports from two runs using a Leoceed-21N HFMD and one run using the Leoceed-18N HFMD are shown in table III. Samples (n = 2 to 4) were averaged and reported ± one standard deviation measurement uncertainty.

**TABLE III.** Blood gas measurements at inlet and outlet of HFMD at 500 ml/min blood flow rates over three runs. A Leoceed-21N HFMD was used for the first two runs and a Leoceed-18N was used for the third run. Measurements are reported as averages over n samples ± 1 standard deviation uncertainty.

| | $pO_2$<br>(kPa) | $pCO_2$<br>(kPa) | $HbSat$<br>(%) |
| --- | --- | --- | --- |
| **Run 1 (21N)** |  |  |  |
| Hb $10.00 \pm 0.05\text{ g/dl}$ | | | |
| Inlet (n=2) | $4.55 \pm 0.03$ | $11.26 \pm 0.08$ | $37.35 \pm 0.03$ |
| Outlet (n=2) | $15.60 \pm 0.1$ | $7.23 \pm 0.05$ | $94.70 \pm 0.07$ |
| **Run 2 (21N)** |  |  |  |
| Hb $10.67 \pm 0.04\text{ g/dl}$ | | | |
| Inlet (n=3) | $4.87 \pm 0.03$ | $8.82 \pm 0.05$ | $43.73 \pm 0.03$ |
| Outlet (n=4) | $9.70 \pm 0.05$ | $6.92 \pm 0.04$ | $85.75 \pm 0.04$ |
| **Run 3 (18N)** |  |  |  |
| Hb $11.20 \pm 0.04\text{ g/dl}$ | | | |
| Inlet (n=4) | $4.20 \pm 0.02$ | $7.43 \pm 0.04$ | $42.68 \pm 0.02$ |
| Outlet (n=4) | $10.23 \pm 0.05$ | $4.65 \pm 0.02$ | $91.08 \pm 0.05$ |

Measurement uncertainties for the blood gas analyser were approximated as 1% for *pO*_2_, *pCO*_2_ and Hb, and 0.1% for *HbSat* based on reported coefficient of variation data [12], [13]. The high values for *pCO*_2_ are in keeping with known changes in stored blood [14].

**Fig. 2.**
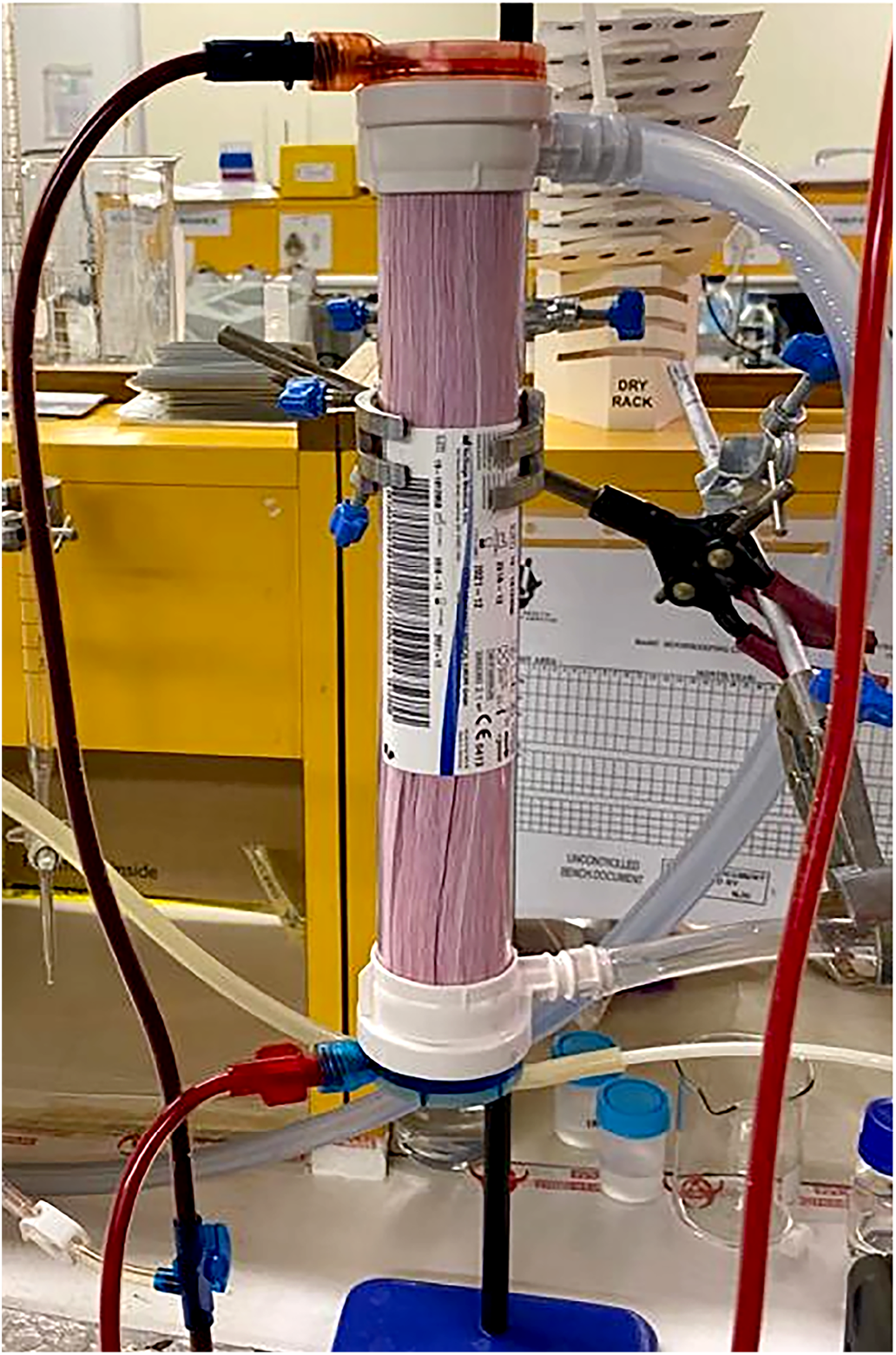
Vertically-mounted Leoceed-21N HFMD showing dark deoxygenated blood entering the device at the top and bright red oxygenated blood exiting from the bottom of the device.

Oxygen content per 100 *ml* of blood (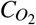) was calculated using a conservative Hufner constant of 1.31 as follows [15]:

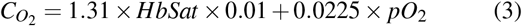

Substituting the differential values for *HbSat* and *pO*_2_ between inlet and outlet into (3), the oxygen transfer rate (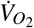) in *ml(STP) · min*^−1^ for a blood flow rate of 500 *ml/min* was calculated as shown in table IV. This table also shows the % of the nominal 250 *ml*(STP) *· min*^−1^ oxygen consumption rate (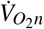) and the % maximum (100 % *HbSat* at outlet at prevailing Hb) oxygen transfer rate (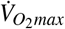).

**TABLE IV.** Oxygen transfer rates (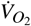) into blood for each of the three runs in *ml · min*^−1^. Also shown are the rounded percentages of the nominal oxygen consumption rates (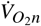) for a typical adult of 250 *ml · min*^−1^ and the maximum oxygen transfer rates (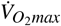) calculated at the prevailing Hb for 100% *HbSat* at the outlet.

| | $\dot{V}_{O_2}$<br>$\text{ml} \cdot \text{min}^{-1}$ | $\% \dot{V}_{O_{2n}}$ | $\% \dot{V}_{O_{2max}}$ |
| --- | --- | --- | --- |
| **Run 1 (21N)** | $38.80 \pm 0.03$ | 16 | 91 |
| **Run 2 (21N)** | $29.70 \pm 0.02$ | 12 | 75 |
| **Run 3 (18N)** | $36.20 \pm 0.03$ | 15 | 85 |

Highest oxygen transfer rates were obtained on Run 1 with the 21N HFMD. Run 2 utilising the same HFMD showed diminished oxygen transfer rates compared to Run 1, probably due to fouling of the HFMD which was noted prior to the second run. Run 3 with the 18N HFMD showed the lowest oxygen transfer rates which is consistent with the smaller surface area.

Using equation (2), mass transfer coefficient × Area product (*k*_0_*A*) estimates for oxygen were 0.72, 0.56 and 0.69. *mg · kPa*^−1^ *· min*^−1^ for runs 1, 2 and 3 respectively. These estimates are in close agreement with theoretical *k*_0_*A* estimates for blood which ranged from 0.389 to 0.787 *mg · kPa*^−1^*min*^−1^. These estimates are also in keeping with minimum estimated *k*_0_*A* of 1.33 *mg · kPa*^−1^*min*^−1^ for minimum oxygen transfer rate of 14.6 *mg · min*^−1^ determined in the water experiments.

### Mass transfer estimates for carbon dioxide

While the principle purpose of this study is blood oxygenation, *CO*_2_ elimination is also an important consideration.

The outlet partial pressure of *CO*_2_ was estimated by performing a *CO*_2_ mass balance using *pCO*_2_ and 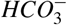, together with the oxygen flow rate of 1250 *ml/min*.

From the *pCO*_2_ and 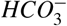 concentrations at the inlet and outlet on each run, and treating *CO*_2_ binding to Hb as negligible, *TotalCO*_2_ concentrations were calculated as shown in table V. Using equation (2), *k*_0_*A* for *CO*_2_ was estimated for runs 1, 2 and 3 as 6.0, 6.6 and 19.4 *mg · kPa*^−1^ *· min*^−1^ respectively. The 3-fold higher value for the 18N dialyser is probably due to the sensitivity of the calculation to the indirectly estimated outlet *pCO*_2_, and uncertainties in the bicarbonate concentrations which are calculated rather than directly measured by the ABL80 FLEX CO-OX.

**TABLE V.** Bicarbonate, *pCO*_2_, and Total *CO*_2_ concentrations at inlet and outlet of HFMD at 500 ml/min blood flow rates over three runs. As outlet gas *pCO*_2_ is approximate, the small uncertainties in measurements are not shown.

| | $HCO_3^-$<br>(kPa) | $pCO_2$<br>(mmol $\cdot l^{-1}$) | Total $CO_2$<br>(mmol $\cdot l^{-1}$) |
| --- | --- | --- | --- |
| **Run 1 (21N)** |  |  |  |
| Inlet (n=2) | 11.62 | 11.26 | 7.10 |
| Outlet (n=2) | 8.75 | 7.23 | 5.21 |
| **Run 2 (21N)** |  |  |  |
| Inlet (n=3) | 10.50 | 8.82 | 6.26 |
| Outlet (n=4) | 8.68 | 6.92 | 5.13 |
| **Run 3 (18N)** |  |  |  |
| Inlet (n=4) | 10.03 | 7.43 | 5.87 |
| Outlet (n=4) | 7.08 | 4.65 | 4.07 |

## IV. DISCUSSION

The minimum membrane oxygen transfer rate in the water experiments informed the decision to proceed to studies with blood.

The experiments with blood flowing at 500 *ml/min* yielded *k*_0_*A* values for oxygen which were in close agreement with the theoretically determined values and the measured value in the water study. This suggests that a Leoceed-21N HFMD would facilitate oxygen transfer rates of about 15% of the nominal ml(STP)/min adult oxygen consumption rate. Using two 21N dialysers in parallel with total flow blood rates of up to a 1 *l/min* would facilitate approximately 30 % of this nominal oxygen consumption rate.

As these studies were performed at ambient pressures (85 *kPa*), operating at sea level and/or designing the system for increased pressure on the inlet gas may achieve an oxygen flux closer to 20% (40% with two HFMDs in parallel) of the nominal adult oxygen consumption subject to sufficient Hb in the blood.

Estimated *k*_0_*A* values for *CO*_2_ were at least an order of magnitude higher than the *k*_0_*A* for oxygen, suggesting that oxygenation will be rate limiting and *CO*_2_ elimination will be readily achieved [16]. *CO*_2_ elimination may even exceed its production rate, possibly requiring reduction of the fresh gas flow rate or adding *CO*_2_ to the fresh gas supply to limit *CO*_2_ losses.

## V. CONCLUSION

A suitable repurposed HFMD with veno-venous access may provide a cost-effective, practical adjunct to respiratory support where ECMO is precluded.

Improvements in performance may be achievable in desperate situations by exceeding the specified maximum blood flow of 500*ml/min*, and increasing the gas-side pressure. However, this should be weighed against risk of damage to the unit resulting in patient harm, including air embolism.

Other considerations such as red blood cell damage, hemodynamic instability, cytokine activation, white blood cell depletion, coagulation and bleeding are likely to be similar to those encountered in dialysis and will need to be addressed by clinicians before considering such an approach.

Dialysis pumps are costly, and presumably most are in service for renal therapy, thus, alternative low-cost pumps would be needed. However, the absence of automatic monitoring may increase risks such as undetected venous air embolisation, and will require weighing the trade-offs in the face of a pandemic.

The results of this study are restricted to the specific devices tested and can not be generalised to other HFMD manufacturers and membranes. Moreover, while no transmembrane leakage was seen on short-term use, further tests are essential to determine the duration that these and other HFMDs can be operated before significant leakage occurs.

Clinical implementation would also require consideration of the placement of the venous catheters, double v.s. single lumens, and the need for non-collapsible catheters under higher flow rates.

This study points to the potential for HFMD repurposing as off-label blood oxygenators in dire, resource-constrained environments. Translational research with full cooperation of regulatory authorities in various jurisdictions would be needed to assess clinical utility, feasibility and safety prior to any consideration of clinical implementation.

## Data Availability

All relevant data is included in the manuscript.

## Acknowledgements

We are grateful to the following people who provided assistance in many different ways to bring this project to fruition: Prof Martin Veller, Prof Ian Jandrell, Dr Paul Freinkel, Mr Ravi Reddy, Mr Andrew Saville, Ms Dolly Mazwi, Mr Humphrey Mbatha, Mr David Heisi, Mr Lebohang Koloti and Mr Liberty Mguni. We thank Fresenius Kabi for donating the dialysis tubing.

